# Plasma pharmacokinetics of high dose oral versus intravenous rifampicin in patients with tuberculous meningitis: a randomized controlled trial

**DOI:** 10.1101/2021.02.11.21250624

**Authors:** Sean Wasserman, Angharad Davis, Cari Stek, Maxwell Chirehwa, Stephani Botha, Remy Daroowala, Marise Bremer, Mpumi Maxebengula, Sonya Koekemoer, Rene Goliath, Amanda Jackson, Thomas Crede, Jonathan Naude, Patryk Szymanski, Yakoob Vallie, Muhammed S Moosa, Lubbe Wiesner, John Black, Graeme Meintjes, Gary Maartens, Robert J Wilkinson

## Abstract

**Background:** Higher doses of intravenous rifampicin may improve outcomes in tuberculous meningitis but is impractical in high burden settings. We hypothesized that plasma rifampicin exposures would be similar between oral 35 mg/kg and intravenous 20 mg/kg, which has been proposed for efficacy trials in tuberculous meningitis.

**Materials and methods:** We performed a randomized parallel group pharmacokinetic study nested within a clinical trial of intensified antimicrobial therapy for tuberculous meningitis. HIV-positive participants with tuberculous meningitis were recruited from South African hospitals and randomized to one of three rifampicin dosing groups: standard (oral 10 mg/kg), high dose (oral 35 mg/kg), and intravenous (intravenous 20 mg/kg). Intensive pharmacokinetic sampling was done on day 3. Data were described using non-compartmental analysis and exposures compared by geometric mean ratio (GMR).

**Results:** Forty-six participants underwent pharmacokinetic sampling (standard dose, n = 17; high dose oral, n= 15; IV, n = 14). Median CD4 count was 130 cells/mm^3^ (IQR 66 −253). Geometric mean AUC_0-∞_ was 47.7 µg·h/mL (90% CI, 33.2 – 68.5) for standard dose; 322.3 µg·h/mL (90% CI,234.6 – 442.7) for high dose; and 214.6 µg·h/mL (90% CI, 176.2 – 261.2) for intravenous. High dose oral dosing achieved higher rifampicin exposure than intravenous: AUC_0-∞_ GMR 0.67 (90% CI, 0.46 −1.0); however, C_max_ GMR was 1.11 (90% CI, 0.81 – 1.59), suggesting equivalence.

**Conclusions:** Plasma rifampicin exposure was similar with high dose oral and intravenous administration. Findings support oral rifampicin dosing in future tuberculous meningitis trials.

## INTRODUCTION

Tuberculous meningitis (TBM) in HIV-positive people carries a mortality approaching 60% (1, 2), and despite antituberculosis therapy, half of all survivors suffer significant neurological sequelae (3). One strategy to potentially improve outcomes is enhanced bacterial killing through optimized antibiotic therapy (4).

Rifampicin is the key agent in TBM therapy; its exclusion from treatment worsens outcomes, and there is high mortality from rifampicin-resistant TBM (5). However, rifampicin is highly protein-bound (6) and the cerebrospinal (CSF) penetration of total drug is poor (7). Standard doses (10 mg/kg) achieve concentrations at only 10-20% of plasma, rarely exceeding the minimum inhibitory concentration of *M. tuberculosis* (8-10). Studies in pulmonary TB have shown that bactericidal activity is related to rifampicin exposure (11, 12) and that microbiological outcomes are improved at higher doses, up to 35 mg/kg (13, 14). A small randomized controlled trial showed survival benefit with the use of intravenous rifampicin 13 mg/kg for Indonesian adults with TBM (15), which had equivalent plasma exposures to oral rifampicin 20 mg/kg (16), A modestly increased oral rifampicin dose of 15 mg/kg did not improve survival in a phase 3 trial (2), however, higher doses may be required to improve outcomes.

Several clinical trials (NCT04145258, ISRCTN42218549, NCT03537495) are currently investigating the safety and efficacy of oral rifampicin doses up to 35 mg/kg for TBM. Because rifampicin has dose-dependent bioavailability (17), and exhibits nonlinear increases in exposure with higher doses (12, 18, 19) 35 mg/kg orally may attain or even exceed intravenous plasma exposures at doses higher than 13 mg/kg. Existing population pharmacokinetic (PK) models can predict plasma rifampicin concentrations at doses up to 40mg/kg orally (20), but this has not been done for intravenous administration where exposure is unaffected by the pre-hepatic first-pass effect (20). This knowledge gap has important implications for TBM trials and the ultimate deployment of intensified antimicrobial therapy for TBM in resource limited settings as intravenous rifampicin has limited availability and use will be associated with increased cost, hospitalization, and complications relating to peripheral venous catherization.

Based on existing PK models of rifampicin (18, 20) and data showing equivalent AUC between 13 mg/kg given intravenously and 20 mg/kg given orally (16), we hypothesized that plasma rifampicin exposures will be similar between oral 35 mg/kg and intravenous 20 mg/kg, which has been proposed for efficacy trials in TBM. To test this, we performed a randomized parallel group PK study nested within a clinical trial of high dose rifampicin for HIV-associated TBM.

## RESULTS

### Participants

Forty-nine participants were enrolled into the parent trial, but 2 participants died and 1 was withdrawn due to late exclusion (eGFR > 20 ml/min) prior to receiving investigational product: 46 participants underwent intensive PK sampling and were included in this analysis (Figure 1).

**Figure 1.**
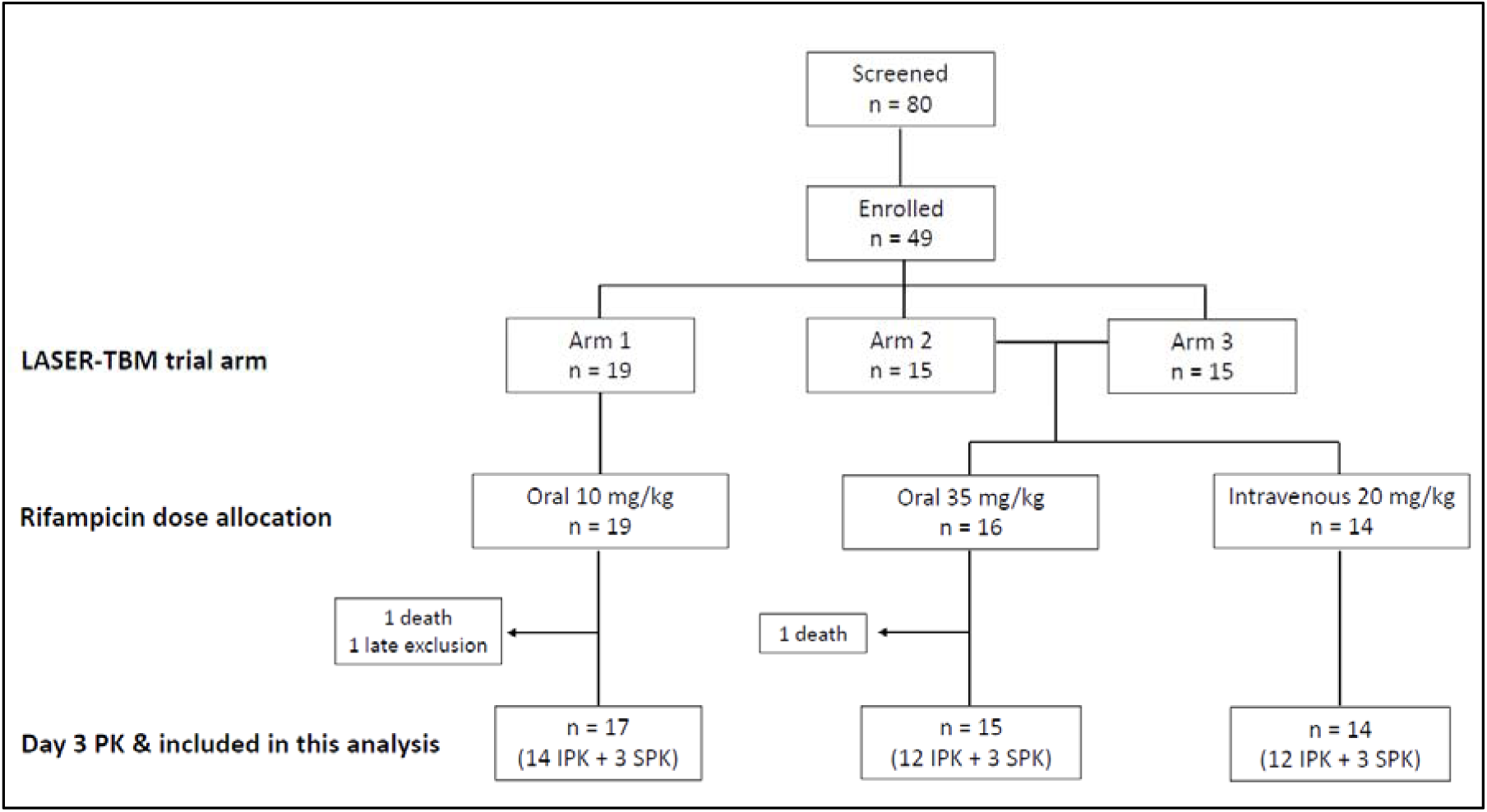
Trial consort. Arm 1, standard TB therapy; Arm 2, high dose rifampicin plus linezolid; Arm 3, high dose rifampicin plus linezolid, plus aspirin; IPK, intensive PK; SPK, sparse PK

Baseline characteristics were well-balanced across rifampicin dosing groups (Table 1). A third of participants had definite TBM, the majority (61%) with MRC Grade 1 disease. Median duration of antituberculosis therapy before the PK visit was 5 days (IQR 4 −6) and was similar across arms (although the PK visit occurred on study Day 2 or 3, up to five days’ standard TB treatment was allowed prior to enrolment). Rifampicin was crushed and administered by syringe for 6 participants (2 high dose group, 4 standard dose group). The duration of intravenous infusion was 60 minutes for all participants except two (15 minutes and 68 minutes).

**Table 1.**
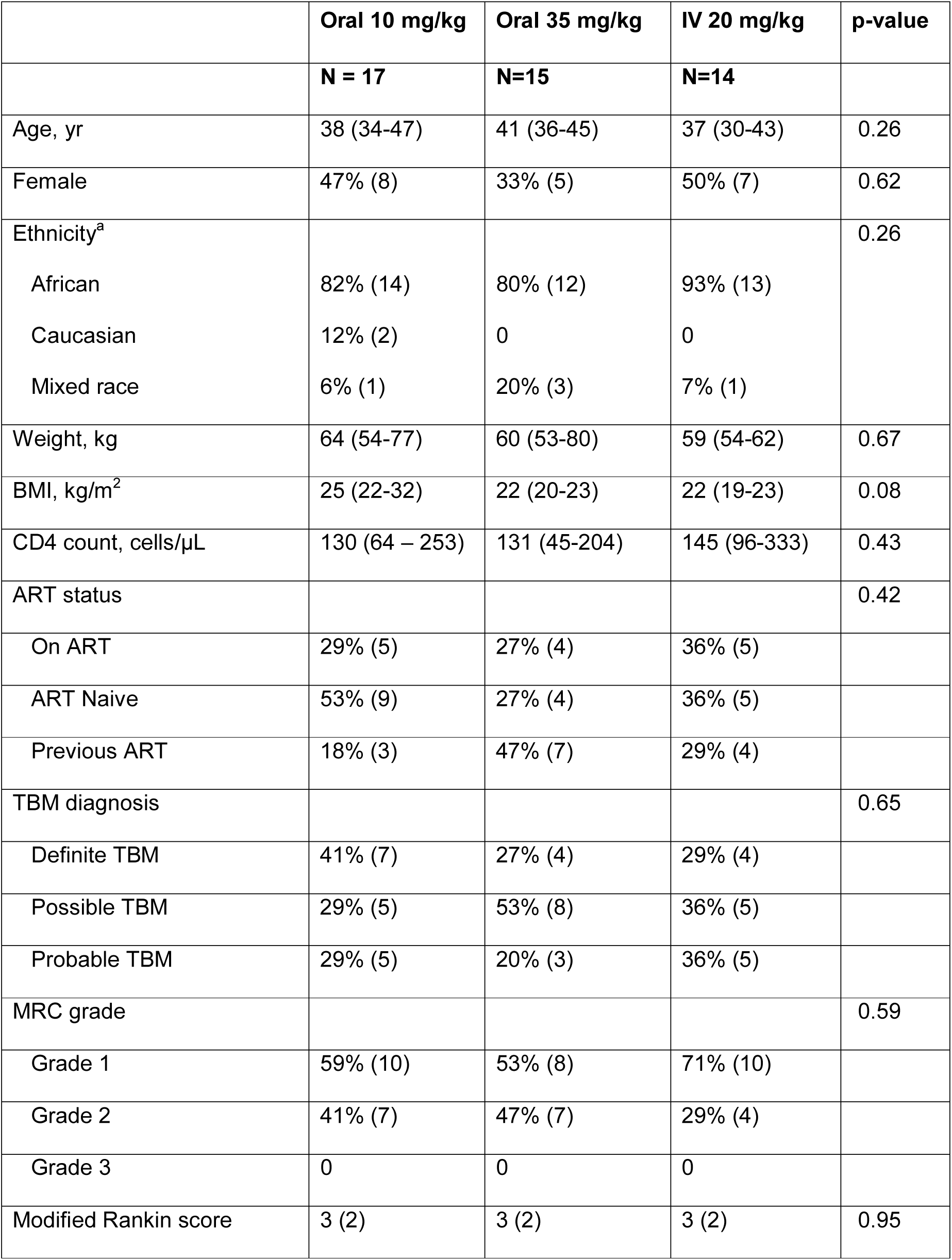
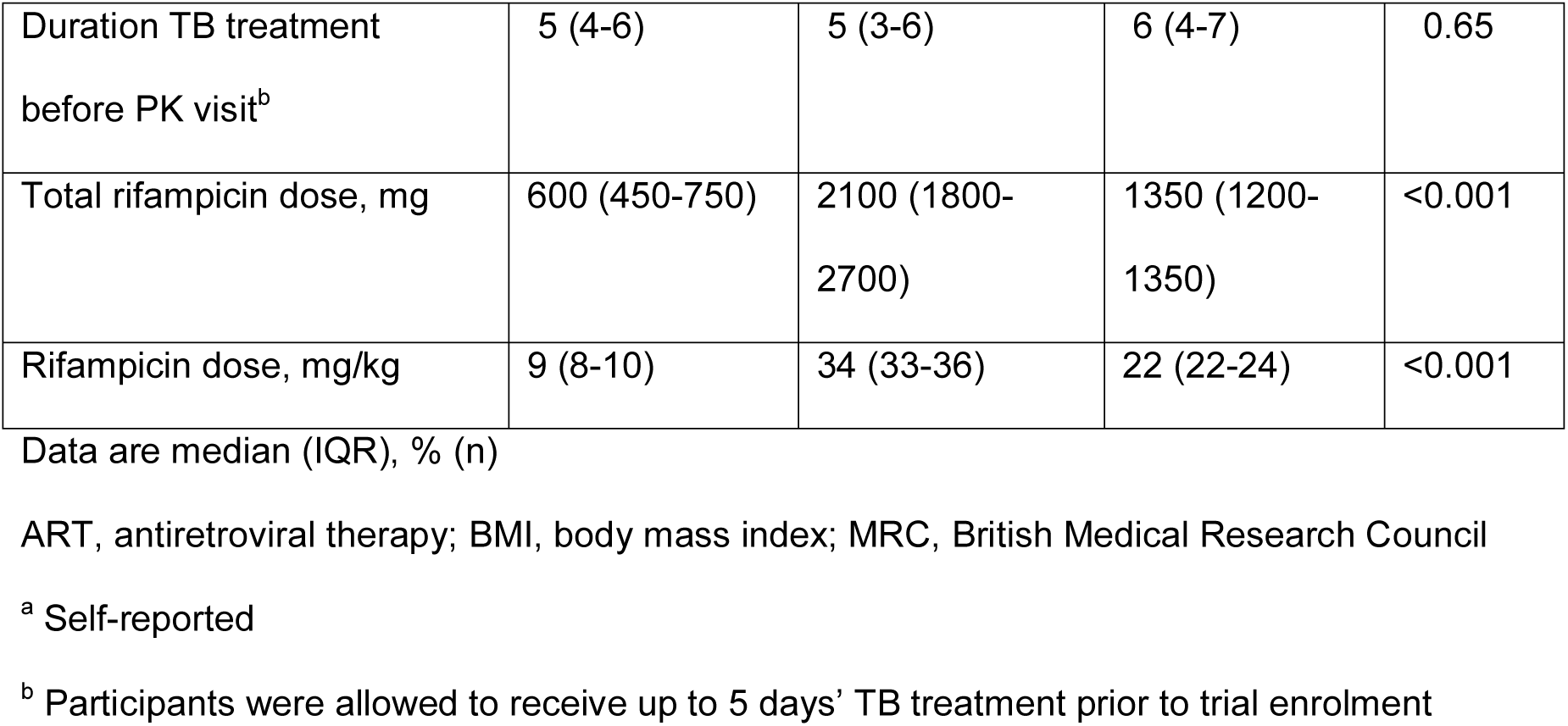
Baseline characteristics.

### PK data

There was a total of 304 PK observations, 40 of which were below the limit of quantification (BLQ). There were 31 full PK profiles after imputation: 10 in the standard dose group, 9 in the high dose oral group, and 12 in the IV group. Trough concentrations were imputed for 9 participants, due to missing 24-hour concentrations in 8 and dosing prior to 24-hour concentration in 1. Pre-dose concentration was imputed for a single participant because of late dosing the day before the PK visit.

Concentration-time profiles in Figure 2 demonstrate much higher concentrations in high dose and IV groups compared with standard dosing. There was high inter-individual variability in plasma concentrations, particularly in the oral dosing groups (standard dose C_max_ %CV 52; high dose oral %CV 48; IV %CV 38), which also showed delayed peaks compared with intravenous administration.

**Figure 2.**
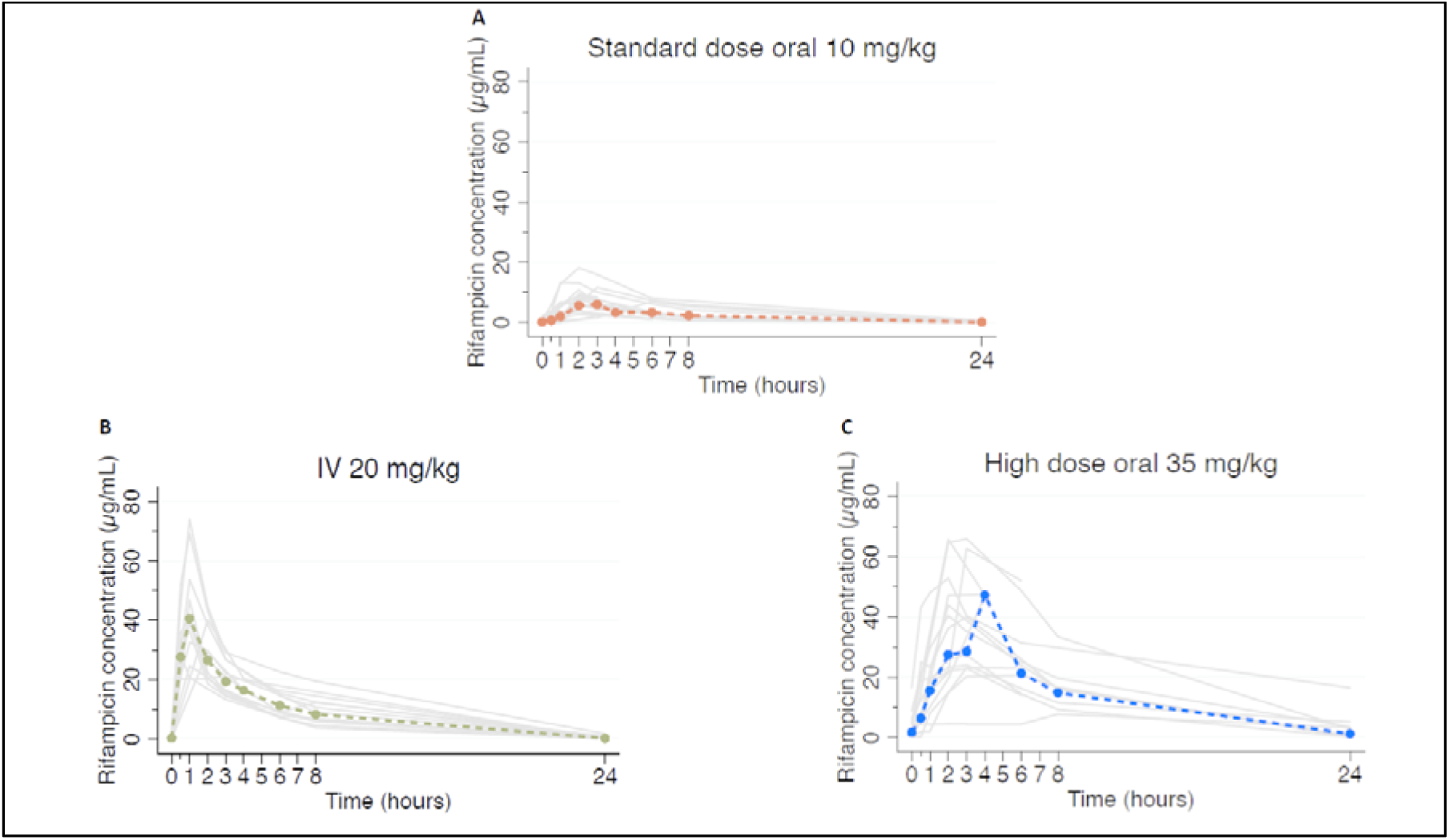
Individual concentration-time profiles. PK profiles for all participants by rifampicin dose allocation. Grey lines indicate individual profiles, coloured dashed lines indicate geometric means.

Table 2 summarizes the estimated PK parameters from observed rifampicin concentrations, by dosing groups. Geometric mean AUC_0-∞_ was 6.8-fold higher for high dose compared with standard dose rifampicin group (ANOVA p < 0.001) but was not significantly different between high dose oral and IV administration (p = 0.96). The lowest AUC_0-∞_ in the high dose oral group (151.9 µg·h/mL) was 2.5-fold higher than the geometric mean AUC in the standard dose group (47.7 µg·h/mL). Geometric mean C_max_ was 4.8-fold higher for high dose oral compared with standard dose rifampicin groups (ANOVA p < 0.001), but similar between high dose oral and IV (p = 0.28). Comparison of exposures across dosing groups is shown in Figure 3. T_max_ was shorter in among those in the IV group: median 1 hour (range 0.5 −2) versus 3 hours (range 2 −8) for high dose oral and 2 hours (range 1 −6) for standard dose. Clearance was significantly higher in the standard dose group (geometric mean 12.6 L/h; range, 4.9 – 53.2) compared with high dose oral (geometric mean 6.8 L/h; range, 2.2 – 16.6) and IV (geometric mean 6.3 L/h; range, 3.9 – 10.7).

**Table 2.** Summary of PK parameters.

| Parameter | Standard dose<br>oral | High dose oral (35<br>mg/kg) | IV (20 mg/kg) | P-value |
| --- | --- | --- | --- | --- |
| AUC, µg·h/mL |  |  |  | < 0.001<sup>a</sup> |
| GM (90% CI) | 47.7 (33.2 – 68.5)* | 322.3 (234.6 – 442.7) | 214.6 (176.2 – 261.2) |  |
| Range | 14.1 - 152.6 | 151.9 - 802.2 | 111.8 - 428.1 |  |
| C<sub>max</sub>, µg/mL |  |  |  | < 0.001<sup>a</sup> |
| GM (90% CI) | 6.9 (5.4 – 8.7)* | 34.7 (26.6 – 45.2) | 38.6 (32.4 – 45.8) |  |
| Range | 2.4 - 18.1 | 7.7 - 66.0 | 20.2 - 74.0 |  |
| T<sub>max</sub>, h |  |  |  | < 0.001<sup>b</sup> |
| Median (range) | 2 (1 – 6) | 3 (2 – 8) | 1 (0.5 – 2)* |  |
| Half-life, h |  |  |  | 0.19<sup>b</sup> |
| Median (range) | 3.2 (2.6 – 6.7) | 3.3 (2.1 – 6.3) | 2.6 (2.2 – 5.4) |  |
| CL, L/h |  |  |  | 0.004<sup>a</sup> |
| GM (90% CI) | 12.6 (8.8 - 18.0)* | 6.8 (4.8 - 9.5) | 6.3 (5.2 - 7.6) |  |
| Range | 4.9 – 53.2 | 2.2 – 16.6 | 3.9 – 10.7 |  |
| %CV | 83.6% | 54.4% | 37.6 |  |
| Vd, L |  |  |  | 0.01<sup>a</sup> |
| GM (90% CI) | 72.9 (42.1 - 126.3)* | 55.2 (30.2 - 100.9) | 27.8 (21.3 - 36.1) |  |
| Range | 23.6 – 191.8 | 21.2 – 116.7 | 13 – 84.3 |  |
| %CV | 184.2% | 150.9% | 59.8% |  |

GM, geometric mean; %CV, coefficient of variation
<sup>a</sup> ANOVA after log transformation
<sup>b</sup> Kruskal-Wallis
\* comparator

**Figure 3.**
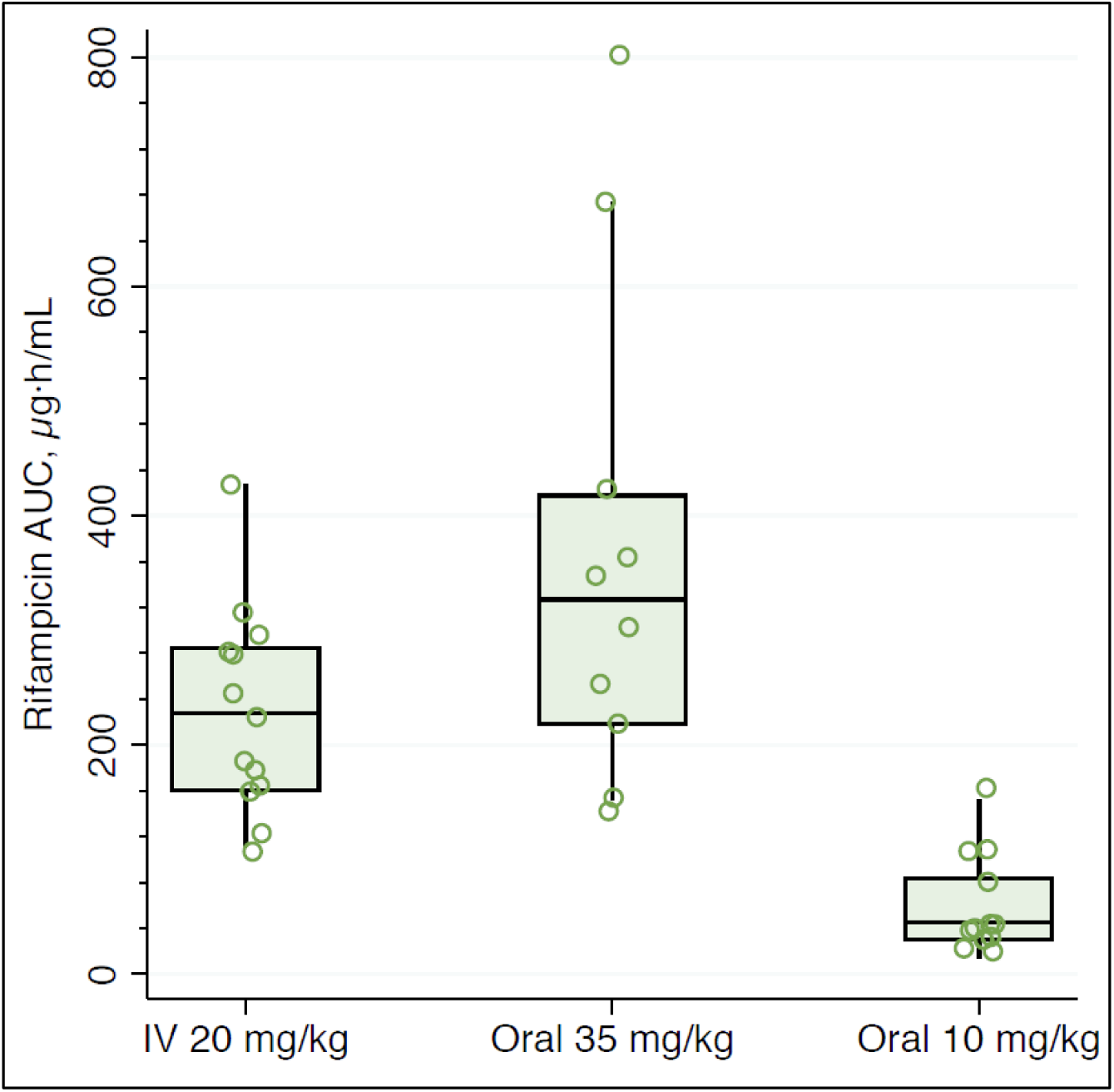
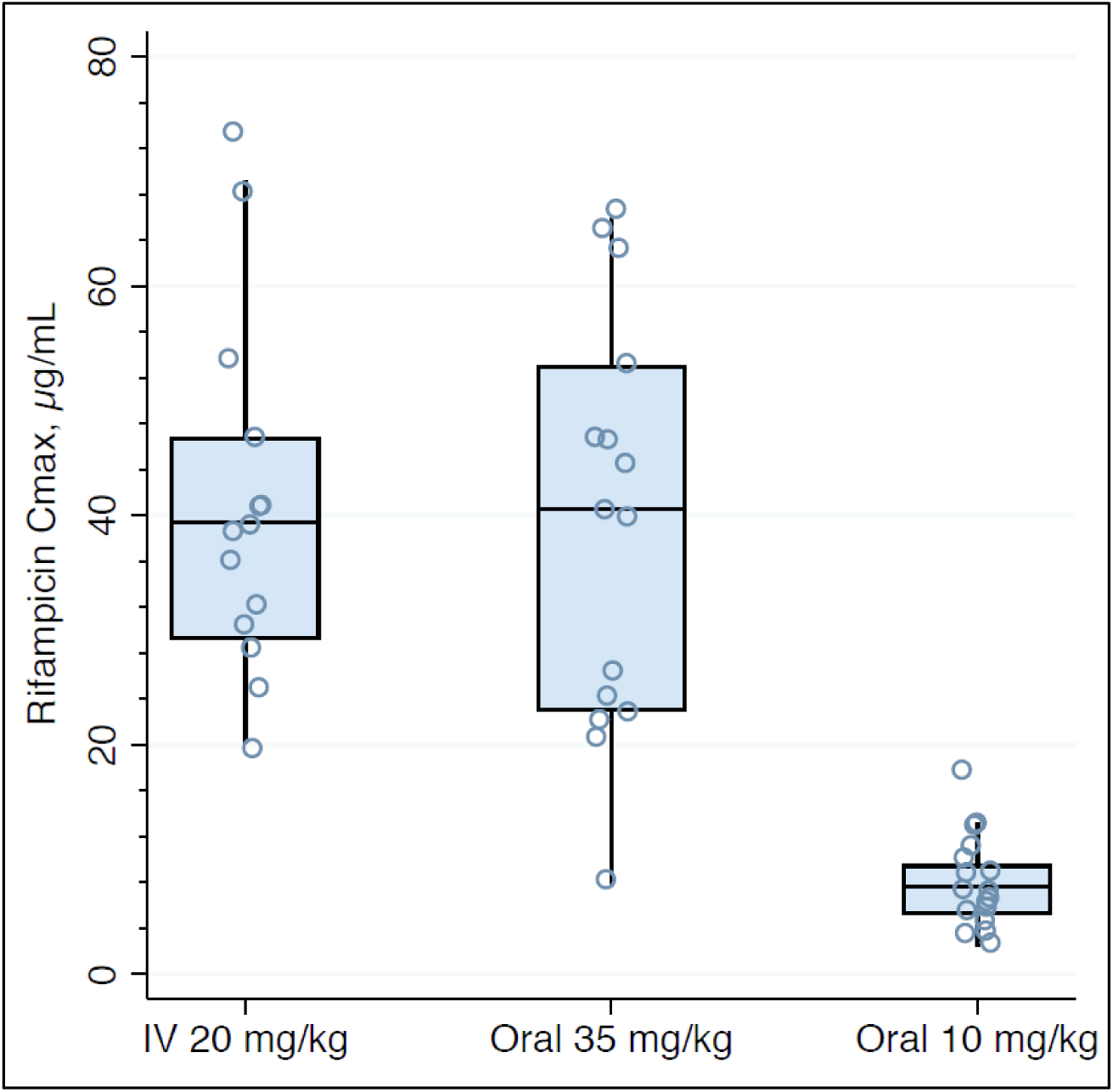
Comparison of exposures across dosing groups. Open circles are individual values for AUC (Figure 3A) and C_max_ (Figure 3B), boxes indicate median and interquartile ranges, whiskers indicate upper adjacent value (1.5x IQR).

In a bioequivalence analysis comparing plasma exposures of high dose oral and IV rifampicin, AUC_0-∞_ GMR was 0.67 (90% CI, 0.46 −1.0), suggesting inequivalence favoring oral dosing; C_max_ GMR was 1.11 (90% CI, 0.81 – 1.59), suggesting equivalence (Figure 4).

**Figure 4.**
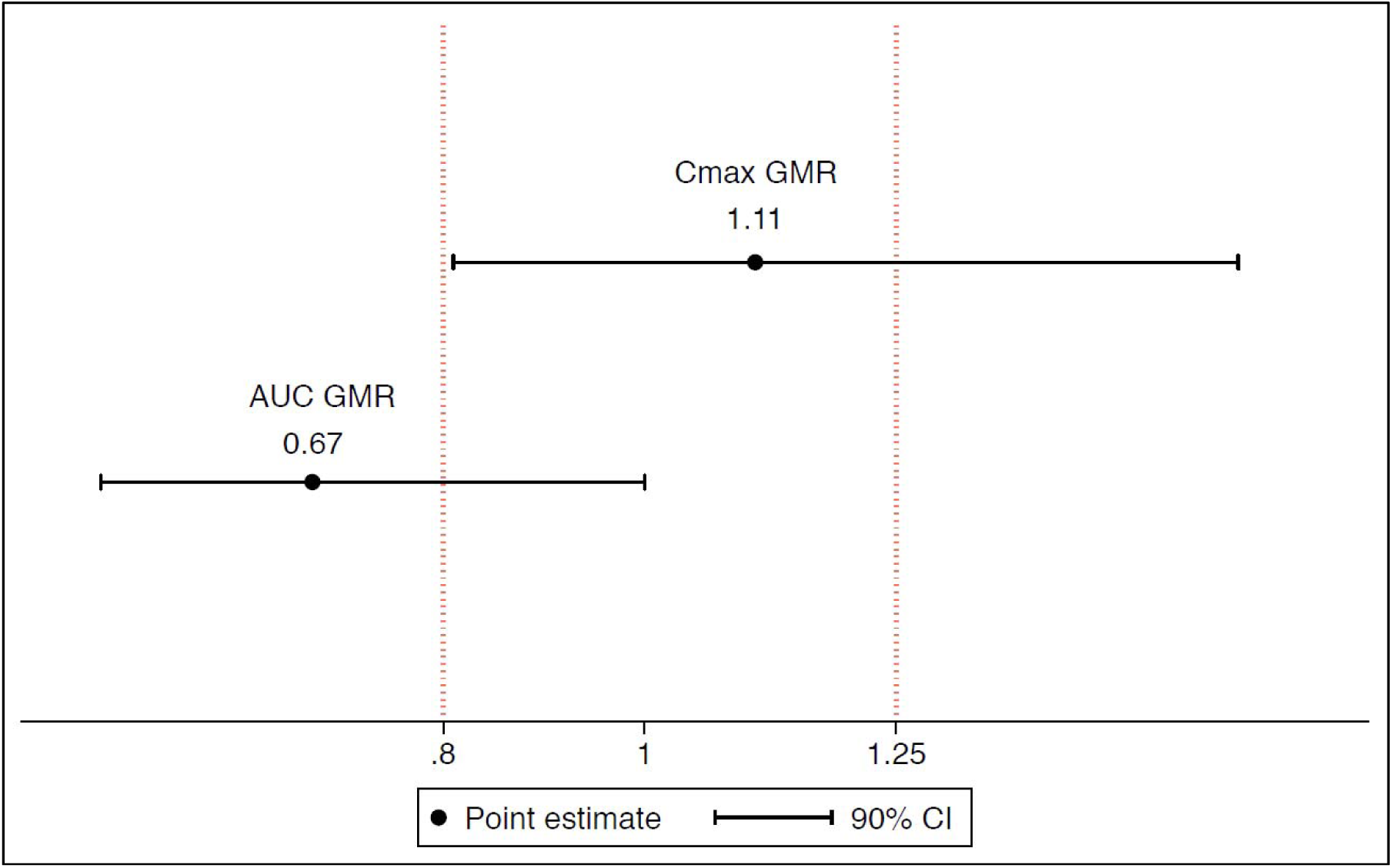
Bioequivalence plot. Point estimates of geometric mean ratios (GMR) for AUC and C_max_, with 90% confidence intervals, with vertical lines indicting bioequivalence margins. The reference measure is intravenous administration (û_IV_/û_oral_), therefore a value > 1 favours intravenous dosing.

Exposures, measured by AUC_0-∞_, were not significantly different across weight bands for the high oral dose (ANOVA p = 0.44), although this had poor precision because the number of participants in each band was small (Figure 5). In an exploratory analysis, exposures were similar after administration of crushed rifampicin via syringe for both the high dose (geometric mean AUC_0-∞_ 383.2 µg·h/mL; n = 2) and standard dose (geometric mean AUC_0-∞_ 38.9 µg·h/mL; n = 4) compared with those who swallowed whole tablets (supplement Figure S2).

**Figure 5.**
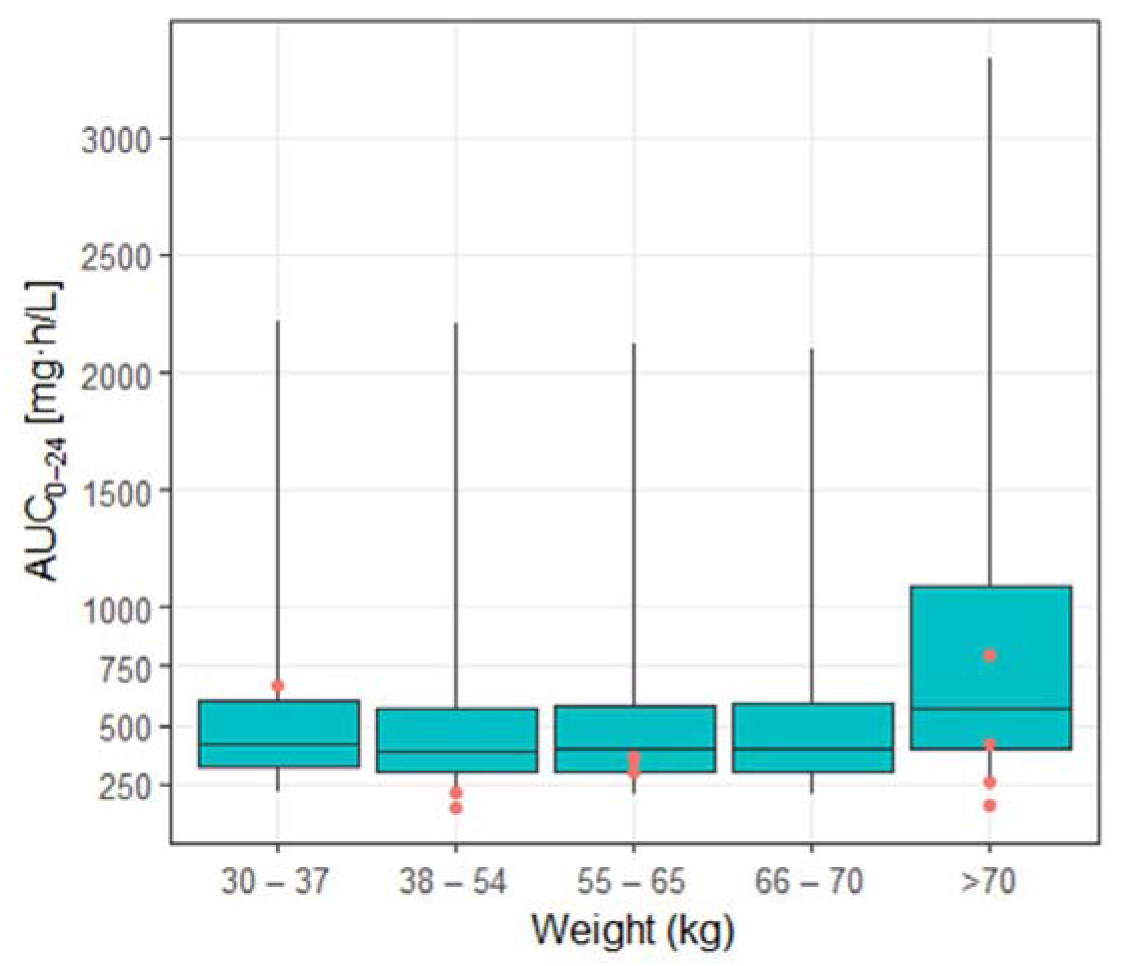
Simulated exposures across LASER-TBM weight bands for 35 mg/kg dosing, with observed exposures superimposed. Boxes indicate median and interquartile range and whiskers indicate range for simulated exposures derived from external cohorts, as described in the text. Red circles indicate observed exposures from the LASER-TBM cohort.

## DISCUSSION

In our randomized controlled trial of South African adults with HIV-associated TBM, plasma rifampicin exposures were similar after an oral 35 mg/kg dose or an intravenous 20 mg/kg dose over the first few days of TB treatment. Consistent with previous studies in both TBM (21) and pulmonary TB (11, 12, 18), there was a non-linear dose-exposure relationship, with higher oral doses achieving supra-proportional increases in exposures compared with standard oral dosing at 10 mg/kg.

The PK efficacy target for rifampicin in TBM is unknown, but it is plausible that dose optimization may lead to improved outcomes. Two small trials conducted in Indonesia suggested a survival benefit with the use of higher oral rifampicin doses up to 30 mg/kg (equivalent to 1,350 mg in that population), and a significant and large effect with the use of intravenous dosing at 13 mg/kg (600 mg) (15, 21). A model-based meta-analysis of those data showed that rifampicin 20 mg/kg given orally resulted in similar exposures to 13 mg/kg given intravenously, and that this translated into a similar effect on TBM survival (22). That same analysis demonstrated an exposure-response relationship and that effect was driven by plasma AUC, similar to the microbiological response in phase 2b pulmonary TB studies (11, 12). Taken together, these findings suggest outcomes in TBM can be improved with use of higher rifampicin doses, and that this is related to overall exposure, irrespective of route of administration.

In our study, geometric mean AUC and C_max_ in the high dose oral and intravenous groups were similar to those reported in other populations (11, 18) and exceeded putative efficacy targets for TBM mortality (estimated AUC 203 µg·h/mL(22) and C_max_ 22 µg/mL (23)). Rifampicin exposures predictably decline at steady-state due to autoinduction and enhanced clearance with repeated dosing (18). Our study was designed to characterize rifampicin PK during the early phase of treatment with the assumption that optimizing exposures would be most critical for anti-mycobacterial effect in this period. Although PK sampling occurred within the first three days of enrolment, median time on rifampicin was 5 days at the time of the PK visit, when substantial autoinduction is expected to have occurred (20). Oral 35 mg/kg dosing would achieve even higher exposures at the start of therapy. In our bioequivalence analysis geometric mean AUC was ∼30% lower with intravenous 20 mg/kg versus oral 35 mg/kg administration, which could be explained by saturation of a first-pass effect at higher oral doses that would not apply to intravenous administration, resulting in a larger reduction in clearance and resultant non-linear dose-exposure relationship with oral dosing, particularly early in therapy. Higher clearance observed in the standard oral dose group supports this, as there is a much lower AUC relative to dose (CL ∝ dose/AUC). As expected, time to maximal concentration was shorter with intravenous administration, but C_max_ was similar to oral dosing at 35 mg/kg. An association between plasma rifampicin C_max_ and survival was found in a small Indonesian TBM study (23) but was not reproduced in a larger Vietnamese trial (24) or in the pooled model-based analysis (22). More rapid intravenous infusion could result in higher C_max_,(25) but the safety and efficacy of this is not established and does not currently justify risks associated with venous catheterisation.

We found large interindividual variability in rifampicin exposure, most pronounced in oral dosing groups. This is a feature of rifampicin PK and relates to effect of absorption delays on bioavailability and saturable kinetics (18, 20, 26). Although AUC was on average significantly higher with 35 mg/kg oral dosing compared with standard dose, certain patients may not attain optimal exposures even at these higher doses. It was somewhat reassuring that, in our study population, the lowest rifampicin exposure in the 35 mg/kg group still exceeded the geometric mean AUC (and equaled the highest AUC) of the standard dose group, suggesting potential benefit from higher dose rifampicin even in the context of highly variable bioavailability. Weight is an important source of rifampicin PK variability; patients with lower weights have relatively lower exposures for a given dose due to allometric scaling on clearance (27). We attempted to compensate for this by implementing a dosing strategy based on simulations using characteristics of a similar population that predicted equitable exposures for the high dose oral group across modified weight bands. Notwithstanding the low number of participants receiving high dose oral rifampicin in each weight band, exploratory analysis suggested no significant difference in observed exposures, providing partial validation of this approach. Another potential source of PK variability is administration of crushed rifampicin tablets, which may affect dissolution characteristics and absorption (26). This is relevant in TBM where patients frequently have reduced levels of consciousness. Reassuringly, the small group of participants (n = 6) who received crushed rifampicin in our study achieved similar exposures to those swallowing whole tablets in their respective dosing groups; this is corroborated by findings from an Indonesian TBM cohort where 60% of participants were administered rifampicin via nasogastric tube but achieved expected increases in exposure at higher doses (21).

There are important limitations to consider when interpreting our findings. The sample size for evaluation of the primary outcome measure (AUC GMR between high dose oral and intravenous rifampicin, n = 29) was smaller than planned due to slow recruitment in the parent trial. However, in a *post hoc* power calculation using the original assumptions, this sample size would provide ∼80% power to detect a difference in AUC of at least 30%, supporting the reliability of our main finding. It is unlikely that the direction of effect would reverse to favor intravenous dosing, even with a larger sample size. The study was not powered to evaluate the impact of physiological or disease characteristics on PK variability; these analyses were not performed but are well-known for rifampicin in similar populations. We did not measure CSF rifampicin concentrations for this analysis because the primary objective was to compare plasma exposure of intravenous versus oral rifampicin. Several studies have shown correlation between plasma and CSF rifampicin exposure with oral dosing in TBM (15, 21, 24), and it is unlikely that CSF PK would be influenced with intravenous administration. Furthermore, plasma rifampicin exposure may be a better predictor of survival than CSF concentrations in TBM (22).

In summary, we have shown that in a population of African patients with HIV-associated TBM, plasma rifampicin exposure was similar when dosed orally at 35 mg/kg or intravenously at 20 mg/kg. We also developed an empiric weight-based dosing strategy for high dose oral rifampicin, which requires validation in a larger cohort. Our findings support high dose oral rifampicin in future TBM trials.

## MATERIALS AND METHODS

### Parent trial and study population

The parent study, called LASER-TBM, is a parallel group, randomized, multi-arm, open label Phase 2a trial evaluating the safety of enhanced antimicrobial therapy with or without host directed therapy for the treatment of HIV-associated TBM. Adults with confirmed HIV and newly diagnosed TBM (based on consensus definitions (28)) were recruited from four hospitals in Cape Town and Port Elizabeth, South Africa. Exclusion criteria included: receipt of more than 5 days antituberculosis medication; evidence of bacterial or cryptococcal meningitis; severe concurrent uncontrolled opportunistic disease; estimated glomerular filtration rate (eGFR) < 20 ml/min (using the Cockcroft-Gault equation); international normalised ratio (INR) > 1.4; clinical evidence of liver failure or decompensated cirrhosis; haemoglobin < 8.0 g/dL; platelets < 50 x109 /L; neutrophils < 0.5 x 109 cells/L; and grade 3 or more peripheral neuropathy on the Brief Peripheral Neuropathy Score. Pregnancy was allowed if gestational age was less than 17 weeks at enrolment.

Eligible and consenting participants were randomized at a ratio of 1.4:1:1 to either a standard of care control group or one of two experimental arms (relatively more participants were allocated to the control group as higher mortality was anticipated with standard of care). Participants allocated to experimental arms 2 and 3 received additional rifampicin (total oral dose 35 mg/kg/day) plus oral linezolid 1,200 mg daily for the first 28 days, reduced to 600 mg daily for the next 28 days; those randomized to experimental arm 3 also received oral aspirin (1000 mg daily). Study treatment was provided in all arms for 56 days, after which participants were referred back to public sector facilities to complete standard therapy for HIV-associated TBM. All participants received antituberculosis chemotherapy as well as corticosteroids as per South African National TB management guidelines. The primary outcome for LASER-TBM was solicited adverse events and deaths in the experimental arms relative to the standard of care control arm at Month 2; efficacy was a secondary outcome, determined at Months 2 and 6.

### Design of PK study

A nested PK study was performed to compare plasma exposure (AUC and C_max_) of intravenous versus oral rifampicin. All consenting LASER-TBM participants allocated to experimental arms underwent a second randomization at the time of study entry, prior to receipt of study drug, to receive either high dose oral (35 mg/kg, according to weight bands described below) or intravenous (IV, 20 mg/kg) rifampicin for the first 3 days of treatment. After Day 3, all participants in experimental arms continued high dose oral rifampicin until Day 56 (supplement figure S1).

Randomization was done in a 1:1 ratio using an electronic randomization tool, and fully integrated with parent trial procedures. A parallel rather than cross-over design was chosen to remove the influence of rifampicin autoinduction on exposure over time, which increases rapidly over the first days of therapy (20). Due to the nature of the intervention, and because the outcome measure is an objective PK endpoint, allocation of intravenous versus oral rifampicin was unblinded.

Intensive plasma PK sampling took place during hospitalization on a single occasion within the first three days of enrolment. Serial venous blood samples were collected into K3EDTA Vacutainer tubes through a peripheral venous catheter pre-dose, and at 0.5, 1, 2, 3, 6, 8-10, and 24 hours after witnessed drug intake (or the start of IV infusion) and an overnight fast. Samples were centrifuged (1,500 x g for 10 minutes) within 1 hour of collection. At least 1.5 mL of plasma was pipetted into polypropylene tubes and immediately frozen at −80°C. Sparse sampling was performed for participants who declined intensive sampling or in whom this failed. Plasma rifampicin concentrations were determined with a validated liquid chromatography tandem mass spectrometry assay developed at the Division of Clinical Pharmacology, University of Cape Town. The assay was validated over the concentration range of 0.117 to 30.0 μg/mL. The combined accuracy and precision statistics of the limit of quantification, low, medium and high-quality controls (three validation batches, n=18) were between 101% and 107%, and 2.7% and 3.7%, respectively.

Demographic and clinical data were collected from participants at the time of LASER-TBM study entry and at the PK visit. Data included biometrics, CD4 count, ART status, TBM diagnosis (definite, possible, or probable by consensus definition (28)) severity (Grade 1 to 3 by British Medical Research Council score) and functional status (modified Rankin score).

### Rifampicin dosing

Oral rifampicin was provided as part of a fixed dose combination tablet with isoniazid, pyrazinamide, and ethambutol (Rifafour, Sandoz) according to standard WHO weight bands for the standard dose group, with top up of single formulation tablets (Rimactane 150 mg, Sandoz; Eremfat 600 mg, Riemser) for the high dose oral group. For participants unable to swallow whole tablets, the rifampicin was crushed, mixed with sterile water, and administered via a syringe. To account for the effect of allometry on clearance at lower weights, we performed simulations to determine the dose of rifampicin required to achieve the most equitable drug exposures across the weight range 30 to 100 kg. Demographic data of a reference cohort of TB patients (n = 1,225), with or without HIV-1 coinfection, recruited in clinical studies conducted in West African countries and South Africa were used for the simulations (27, 29-31). An additional 12,250 virtual patients were generated using the weight and height distributions of the 1,225 patients to increase the number of patients with a weight close to the boundaries of the weight range. Parameter estimates of a population PK model for rifampicin were used to simulate (100 replicates) rifampicin exposures (18). Four dosing scenarios were evaluated using the weight-band based dosing with 4-drug fixed dose combination (FDC) tablets and extra rifampicin tablets, with each tablet containing 150 mg or 600 mg rifampicin. The FDC tablets were assumed to have 20% reduced bioavailability based on data from a clinical trial where the same formulation was used (32). The weight bands with the most balanced distribution in predicted exposures were used to dose oral rifampicin in the trial (supplement table S1 and figure S2). Intravenous rifampicin (Eremfat 600 mg vials, Riemser) was administered according to weight bands (supplement table S2) as a 1-hour infusion, in accordance with instructions in the package insert, by nursing staff of the parent trial.

### Analysis

The study was powered to detect a difference in exposure between oral and intravenous administration, defined as an AUC geometric mean ratio (GMR) < 0.8.(33) Assuming increased variability with oral dosing (coefficient of variance, %CV 34)(18) versus intravenous dosing (%CV 20), a sample size of 50 participants was planned to provide 80% power to demonstrate this with 90% two-sided confidence.

Demographic and clinical characteristics were summarized and compared using the Wilcoxon rank-sum test for continuous variables and 𝒳^2^ test for dichotomous variables. Non-compartmental analysis was used to estimate rifampicin PK parameters from observed concentrations. The area under the concentration-time curve, extended to infinity (AUC_0-∞_), was calculated as AUC_0-tmax_ (using the trapezoid method) + AUC_tmax-∞_ (estimated by extending the curve with linear fit to the log of the concentration). Trough concentration (Cτ) was defined as the plasma concentration 24 hours after observed intake (actual or imputed, as described in the supplement). The elimination rate constant (k_e_) was assessed by linear regression analysis of the last three concentrations in the terminal log-linear period. The apparent clearance of the drug (CL/F) and the volume of distribution after oral administration (Vd/F) were calculated using standard equations. %CV was calculated as mean/standard deviation x 100. Differences between log-transformed PK parameters across the three study groups were tested by one-way analysis of variance (ANOVA); the Kruskal–Wallis test was used for time to maximal concentration (T_max_) and half-life. The means of log-transformed values for exposure parameters (log-normally distributed) were back-transformed to obtain geometric means; GMR was calculated for AUC and Cτ, with oral administration as the reference (û_IV_/û_oral_). Fieller’s method was used to estimate 90% confidence intervals for GMR. Statistical analysis was performed using Stata version 14.2 (StataCorp).

### Ethics

This research was conducted in accordance with the Declaration of Helsinki and was approved by the University of Cape Town Human Research Ethics Committee (Ref 293/2018) and the Walter Sisulu University Human Research Committee (Ref 012/2019). The parent trial (LASER-TBM) is registered on clinicaltrials.gov (NCT03927313) and approved by the South African Health Products Regulatory Authority (Ref 20180622).

## Data Availability

Original data available on request

## ACKNOWLEDGEMENTS

We thank all study participants and clinical staff for generously contributing their time; our trial nurses, Louise Lai Sai, Vuyiswa Nonkwelo and Thandi Sihoyiya; laboratory staff Nonzwakazi Bangani, Francisco Lakay, and Fatima Abrahams; trial pharmacist Yakub Kadernani; and Celeste Worship for data capturing. We also thank the Clinical Research Centre at the University of Cape Town for pharmacy and other clinical trial support.

SW was supported by the European & Developing Countries Clinical Trials Partnership (Grant number CDF1018), Wellcome Trust (Grant number 203135/Z/16/Z and 104803), and National Institutes of Health (K43TW011421). AGD is supported through a UCL Wellcome Trust PhD Programme for Clinicians Fellowship (award number 175479). GrM was supported by the Wellcome Trust (098316, 214321/Z/18/Z, and 203135/Z/16/Z), and the South African Research Chairs Initiative of the Department of Science and Technology and National Research Foundation (NRF) of South Africa (Grant No 64787). The funders had no role in the study design, data collection, data analysis, data interpretation, or writing of this report. The opinions, findings and conclusions expressed in this manuscript reflect those of the authors alone. RJW receives support from the Francis Crick Institute which is funded by UKRI (FC0010218). He also receives support from Meningitis Now and NIH (R01AJ145436). Research reported in this publication was also supported by National Institute of Allergy and Infectious Diseases (NIAID) of the National Institutes of Health (Award nos. UM1 AI068634, UM1 AI068636 and UM1 AI106701).

No conflicts of interest to declare. SW conceived the study, collected data, did the analysis, wrote the first draft of the manuscript. AD was involved in study inception and led trial implementation and data collection. MC did the rifampicin dosing simulations. SK was the lead trial pharmacist. AJ developed the trial database and oversaw data management. RG was the trial project manager. MM was the study coordinator. LW performed the drug assays. GrM provided scientific input and edited the manuscript. GaM contributed to study conception and design and edited the manuscript. RJW led study inception, funding, supervision, and edited the manuscript. All other authors collected data and reviewed the manuscript.

