## Supplementary material for "Plasma pharmacokinetics of high dose oral versus intravenous rifampicin in patients with tuberculous meningitis: a randomized controlled trial"

Sean Wasserman^1,2#^, Angharad Davis^1,3,4^, Cari Stek^1,5,6^, Maxwell Chirehwa^7^, Stephani Botha^1^, Remy Daroowala^1,5^, Marise Bremer^1,8^, Mpumi Maxebengula^1^, Sonya Koekemoer^1^, Rene Goliath^1^, Amanda Jackson^1^, Thomas Crede^6,9^, Jonathan Naude^6,9^, Patryk Szymanski^6,9^, Yakoob Vallie^6,10^, Muhammed S Moosa^6,10^, Lubbe Wiesner^7^, John Black^8^, Graeme Meintjes^1,2^, Gary Maartens^1,7^, Robert J Wilkinson^1,2,3,4,5^

1. Wellcome Centre for Infectious Diseases Research in Africa, Institute for Infectious Disease and Molecular Medicine, University of Cape Town, Observatory, 7925, South Africa.
2. Division of Infectious Diseases and HIV Medicine, Department of Medicine, University of Cape Town, Observatory, 7925, South Africa.
3. Francis Crick Institute, Midland Road, London, NW11AT, UK.
4. Faculty of Life Sciences, University College London, Gower Street, London, WC1E 6BT, UK.
5. Department of Infectious Diseases, Imperial College, London, W12ONN, UK.
6. Department of Medicine, University of Cape Town, Cape Town, South Africa.
7. Division of Clinical Pharmacology, Department of Medicine, University of Cape Town, Observatory, 7925, South Africa.
8. Livingstone Hospital Complex, Eastern Cape Department of Health, Port Elizabeth, South Africa.
9. Mitchells Plain Hospital, Western Cape Department of Health, Cape Town, South Africa.
10. New Somerset Hospital, Western Cape Department of Health, Cape Town, South Africa.

**IMPUTATION STRATEGY**

Concentration-time profiles were inspected for each participant to compare pre-dose and 24-hour concentrations. In cases where the 24-hour concentration was missing, these were imputed as pre-dose concentrations if two prior observations were available in the elimination phase. The 24-hour concentration was considered highly unlikely to represent the true trough value where it exceeded the pre-dose concentration and was > 50% of the concentration at the prior sampling time point. This was based on the published elimination half-life of rifampicin,^16^ and the assumption that the 24-hour concentration would therefore fall below the 6- or 8-hour concentration in the absence of additional dosing. In these cases, the 24-hour concentration was imputed from the pre-dose concentration. Where the pre-dose concentration exceeded the 24-hour concentration by > 2-fold, indicating late dosing prior to the PK visit, the pre-dose concentration was replaced by C24*K_e_ to adjust for contribution to AUC. Concentrations reported as below the limit of assay quantification (BLQ) were imputed as 50% of the lower limit of detection (i.e. 0.585 µg/mL).

**FIGURES**

**Figure S1. Trial schema**

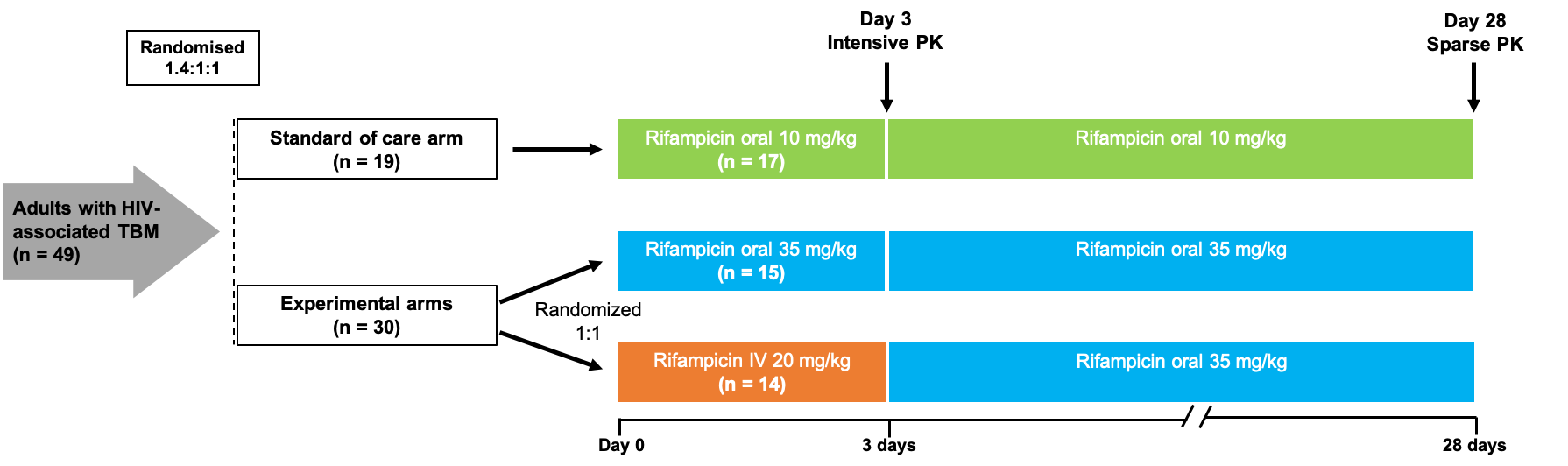

**Figure S2. Simulations showing balanced exposures across weight bands with LASER-TBM dosing table**

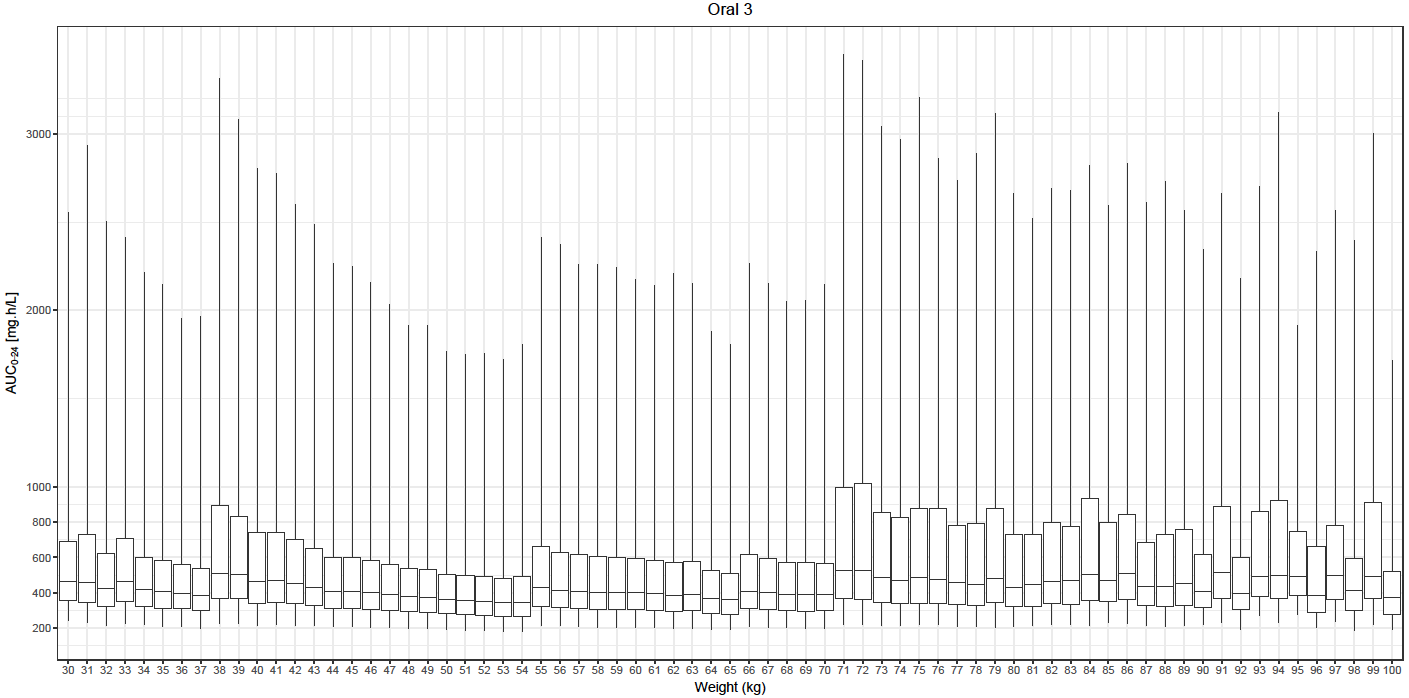

**Figure S3. Exposures from swallowed versus crushed administration of rifampicin tablets for oral 35 mg/kg (Fig. S2A) and 10 mg/kg (Fig. S2B).**

**Fig S3A**

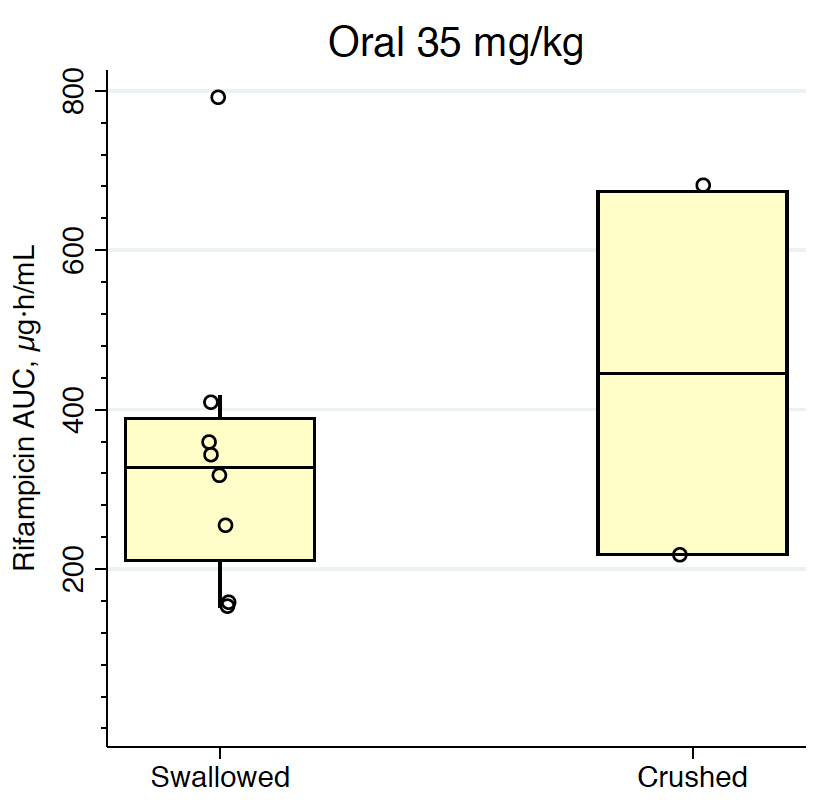

**Fig S3B**

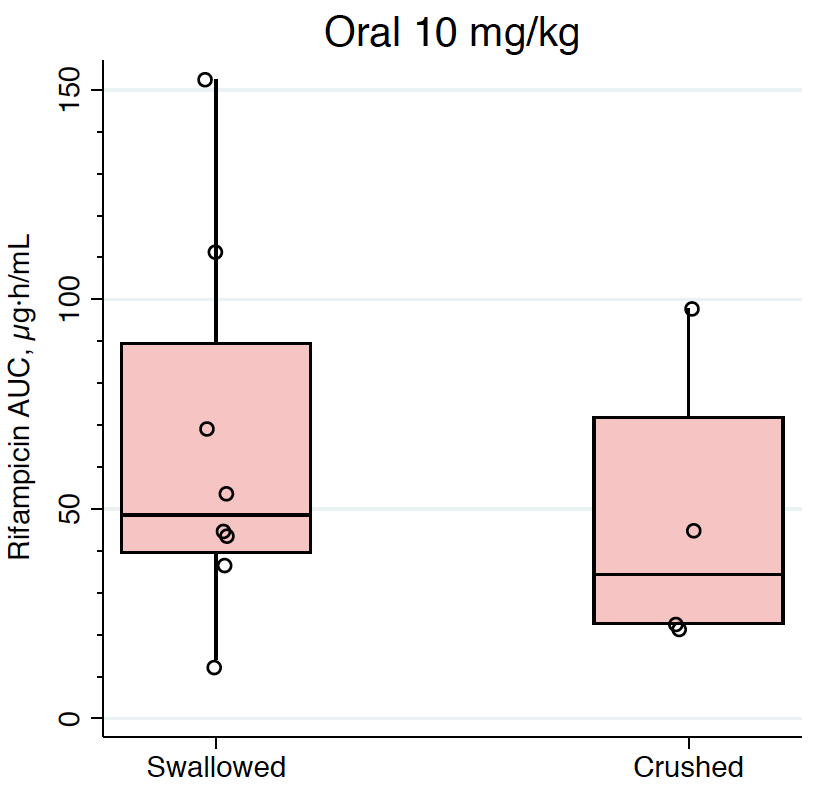

**Figure S4. Exposures from observed data across LASER-TBM weight bands for 35 mg/kg dosing.**

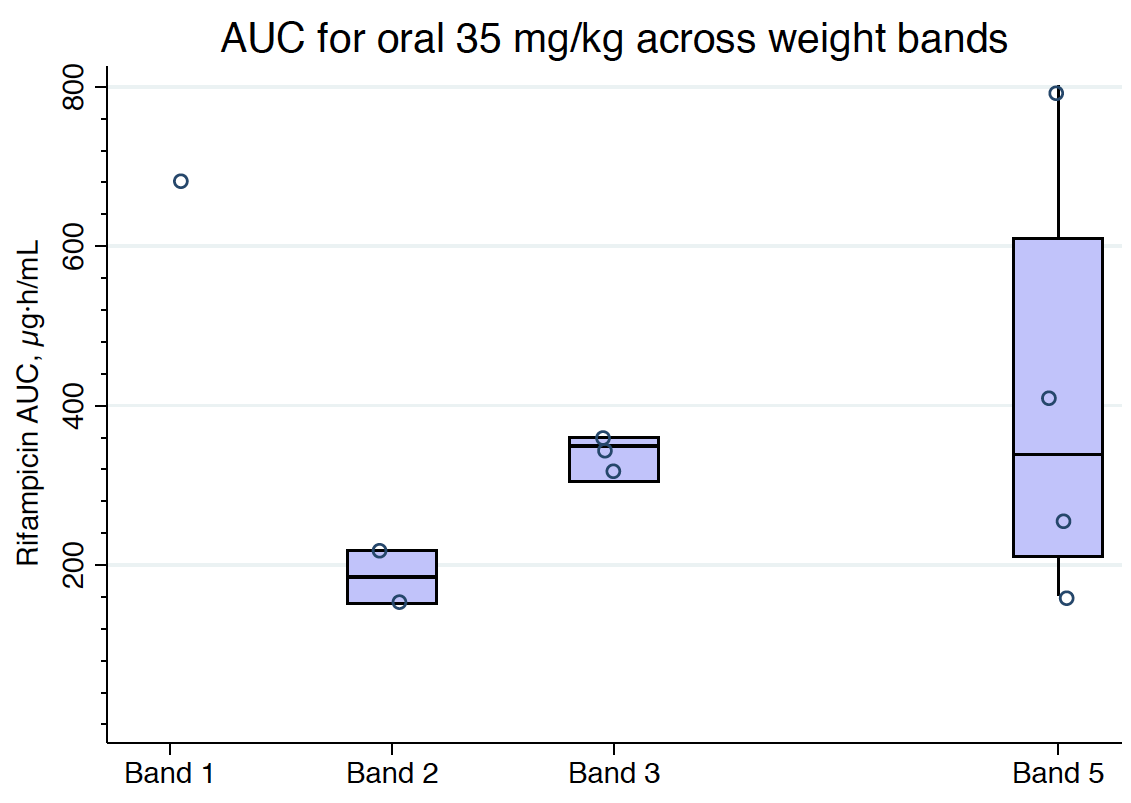

**TABLES**

**Table S1. Weight bands for oral rifampicin dosing**

| **LASER-TBM bands** | **Band 1** | **Band 2** | **Band 3** | **Band 4** | **Band 5** |
| --- | --- | --- | --- | --- | --- |
| **Weight range** | 30 – 37 kg | 38 – 54 kg | 55- 65 kg | 66 - 70 | > 70 kg |
| **R_10_HZE (WHO)** | 300 | 450 | 600 | 600 | 750 |
| **R_25_ additional** | 1200 | 1350 | 1500 | 1650 | 1950 |
| **Total RIF (~35 mg/kg)** | 1500 | 1800 | 2100 | 2250 | 2700 |

**Table S2. Weight bands for intravenous rifampicin dosing**

|  | **Band 1** | **Band 2** | **Band 3** | **Band 4** | **Band 5** | **Band 6** |
| --- | --- | --- | --- | --- | --- | --- |
| Weight range | 30 – 33 kg | 34 - 37 kg | 38 – 54 kg | 55- 65 kg | 66 - 70 kg | > 70 kg |
| HZE tabs | 2 | 2 | 3 | 4 | 4 | 5 |
| R_20_ IV | 900 | 1050 | 1200 | 1350 | 1500 | 1650 |
| Total Rif | 900 | 1050 | 1200 | 1350 | 1500 | 1650 |
